# Advance requests for MAiD in dementia: Policy recommendations emerging from a mixed-methods study of the views of the Canadian public and MAiD practitioners

**DOI:** 10.1101/2021.03.28.21254508

**Authors:** Adrian C. Byram, Ellen R. Wiebe, Sabrina Trembley-Huet, Peter B. Reiner

## Abstract

**Background:** The Canadian public has repeatedly expressed its desire for advance requests for Medical Assistance in Dying (MAiD) after dementia diagnosis, yet MAiD practitioners’ willingness to accede to such advance requests is unknown. This study explores the extent and nature of any gap between the public’s desire for, and practitioners’ willingness to provide MAiD, and identifies policies to ameliorate such a gap.

**Methods:** Two complementary mixed-method surveys gathered data from convenience samples of 83 Canadian adults and 103 MAiD practitioners. The public survey asked participants which of five specific circumstances commonly encountered in dementia they would include in their advance requests. The practitioner survey queried the validation level participants would require before providing MAiD in each specific circumstance. Participants’ reasons were probed using thematic analysis of open-ended questions.

**Results:** On average, 77% of public participants indicated they definitely or probably would include each of the five specific circumstances in their advance requests for MAiD. As validation level decreased from patient consent to patient assent, family assent, or advance request alone, the magnitude of the gap between the public’s desire and practitioners’ willingness increased. The practitioners’ qualitative data contained many practical insights from which emerged seven policy recommendations to ameliorate this gap and increase the likelihood of honouring patient requests.

**Interpretation:** The study provides evidence of a gap between public desire for, and practitioner willingness to provide MAiD in dementia. The policy recommendations are relevant to consideration of legislation for advance requests for MAiD.

## Introduction

Medical Assistance in Dying (MAiD) became legal in Canada in 2016.^1^ The legislation excluded advance requests – statements made while fully competent that memorialize a person’s desire for MAiD at some point in the future when they may have lost decisional capacity. In early 2020, a Department of Justice public consultation survey found that Canadians overwhelmingly support advance requests for MAiD: 79.4% of the >300,000 respondents endorsed the idea that a MAiD practitioner should be allowed to fulfill an advance request for MAiD in the case of dementia.^2^ In March of 2021, the legislation was amended with Bill C-7^3^ which did not legalize advance requests but allowed advance consent: a patient at risk for losing competence can enter into a ‘written arrangement’ with a MAiD provider to initiate MAiD at some future date after the patient is no longer able to consent. In essence, advance consent enables advance requests if a MAiD provider is willing to enter into a written arrangement.^4^

Implementing advance requests or advance consent will be challenging. The Netherlands, where advance requests for euthanasia have been legal since 2002, provides an illuminating example. A review of the actual practice revealed that practitioners rarely comply with dementia patients’ wishes unless some form of assent is obtained.^5^ Thus, there may be a gap between the public’s desire for practitioners to accept advance requests for MAiD and practitioners’ willingness to do so. Our research objectives are to characterize the extent and nature of any such gap in the Canadian context and to identify actionable policies and guidelines that could ameliorate it.

## Methods

This study employed two complementary, mixed-method web-based surveys, the first targeted to adult members of the Canadian public, the second to active MAiD practitioners. Both surveys asked respondents about their reactions to a set of five different specific circumstances (SCs) commonly encountered in dementia that could be included in an advance request to initiate the provision of MAiD even if the patient no longer has decisional capacity:

1. Loss of Personal Dignity: the patient can no longer feed and toilet themself;
2. Loss of Freedom: the patient has to be kept in a locked facility because otherwise they would wander off, endangering themselves;
3. Loss of Ability to Recognize Family: the patient can no longer identify their spouse, siblings, or children by name, even when standing in front of them;
4. Loss of Ability to Form New Memories: the patient can no longer remember even simple new facts for more than 2 minutes; and
5. Loss of Ability to Control Behaviour: the patient requires restraints or daily treatment with drugs to control aggressive or inappropriate behaviour.

The public survey asked respondents whether they would include each of these SCs in their advance request (using a scale of definitely yes, probably yes, probably not, definitely not). They were also asked for their reasons why or why not (free text response), as well as demographic information. The survey also included question D2 of the Department of Justice’s 2020 public consultation.^2^

The MAiD practitioner survey asked respondents whether they would or would not be willing to provide MAiD in response to an advance request that included each of the SCs, given each of the following levels of validation at the time of provision: the patient consents, assents, could not assent but family assents, or the sole direction for initiating MAiD comes from the existence of the advance request itself. MAiD practitioners were then asked their reasons for responding as they did (free text response). They were then shown the reasons given by the public for choosing to include or not include these SCs in their advance requests, and were asked if the public’s reasons were sufficient to provide MAiD, both as a Yes/No and as a free text response. After indicating the number of times they had personally provided MAiD, they were given a final option to express any further insights they may have.

In order to determine the *extent* of any gap between the public and MAiD practitioners, we compared the percentage of the public who indicated that they would definitely or likely include each SC in their advance requests with the percentage of MAiD practitioners who would definitely or likely provide MAiD in a given SC given each of 4 different levels of validation (consent, assent, etc.).

The *nature* of the gap between the public and MAiD practitioners was analyzed in two stages. We first applied thematic analysis^6^ to the public’s free text responses for the reasons why they would or would not include the SCs in their advance requests. The coding of these themes was conducted independently by ACB and PBR, then reconciled and refined until Cohen’s kappa > 0.8. The public’s reasons discovered by this process were then incorporated into the MAiD practitioners’ survey, asking whether the MAiD practitioners found these reasons sufficient for providing MAiD in response to an advance request (both yes/no and free response). In the second stage, ACB and PBR again applied thematic analysis to free text responses of the MAiD practitioners.

In order to identify clinically actionable policies and guidelines, using a multi-round process of coding, reconciliation, and refinement, ACB and PBR transformed the required free-text themes expressed by the MAiD practitioners as to their willingness to provide MAiD for the SCs, as well as their optional responses to their agreement with the public’s reasons for wanting MAiD and, at the end of the survey, a request to share additional insights about MAiD into an interpretive description yielding “tentative truth claims about a clinical phenomenon.”^7^

For the public survey we recruited convenience sample of Canadians (≥ 18-years-old) from a Qualtrics survey panel. Data were collected Aug 18-19, 2020.

For the MAiD practitioners’ survey, we recruited a convenience sample of active MAiD practitioners from the Canadian Association of MAiD Assessors and Providers (CAMAP|ACEPA) and La Communauté de Pratique AMM-Quebec (CPAQ); invitations explaining the purpose of the study with a clickable link were emailed to all 169 CAMAP listserve members and 77 CPAQ listserve members. Data were collected Oct 6-22, 2020 (CAMAP), and Oct 13-23, 2020 (CPAQ). Free-text answers to the French-language survey were translated into English by ST-H and validated by ACB; the English and French language responses were then amalgamated.

This study was approved by the University of British Columbia Research Ethics Board (H20-02437).

## Results

Completed surveys were received from 105 members of the general public and 103 MAiD practitioners (providers and assessors); 22 surveys from the public were rejected because the free-text answers were non-responsive to the question. Public participant demographics are shown in Table 1; prior experience with provision of MAiD by the MAiD practitioners is shown in Table 2.

**Table 1:** Public participant demographics

|  |  |
| --- | --- |
| Gender | 58% female (n=48) |
| Age | Average 45.4 years; Range 18 to 79 years |
| Race | White 83%;<br>East Asian 10%<br>South Asian 4%<br>First Nations 2%<br>Black 1%<br>Other 1% |
| Province | ON 42%, NS 18%, AB 14%, BC 11%, QC 6%, NB 5%, SK 2%, MB 1%<br>all other provinces and territories 0% |

**Table 2:** Practitioners’ experience in providing MAiD

|  |  |
| --- | --- |
| Never provided MAiD (assessors) | 12% |
| Under 10 provisions | 13% |
| 10 to 50 provisions | 43% |
| Over 50 provisions | 33% |

We presented public respondents with question D2 of the Department of Justice’s 2020 public consultation^2^ which asked whether a MAiD practitioner ought to comply with an advance request for MAiD for an individual with dementia once an unspecified ‘specific circumstance’ arises; 86% of the respondents to our survey answered yes, while 79.4% of the respondents to the government survey answered yes.

We hypothesized that asking people to consider an unspecified ‘specific circumstance’ as the event that would set in motion the process of MAiD might overestimate support for advance requests, and that when circumstances were more fully specified, support might diminish. To test this hypothesis, we asked respondents whether they would include in their advance request any of the five SCs described in methods. On average, 77% of respondents indicated that they definitely or probably would include them in their advance requests; no SC garnered less than 70% support (Figure 1).

**Figure 1.**
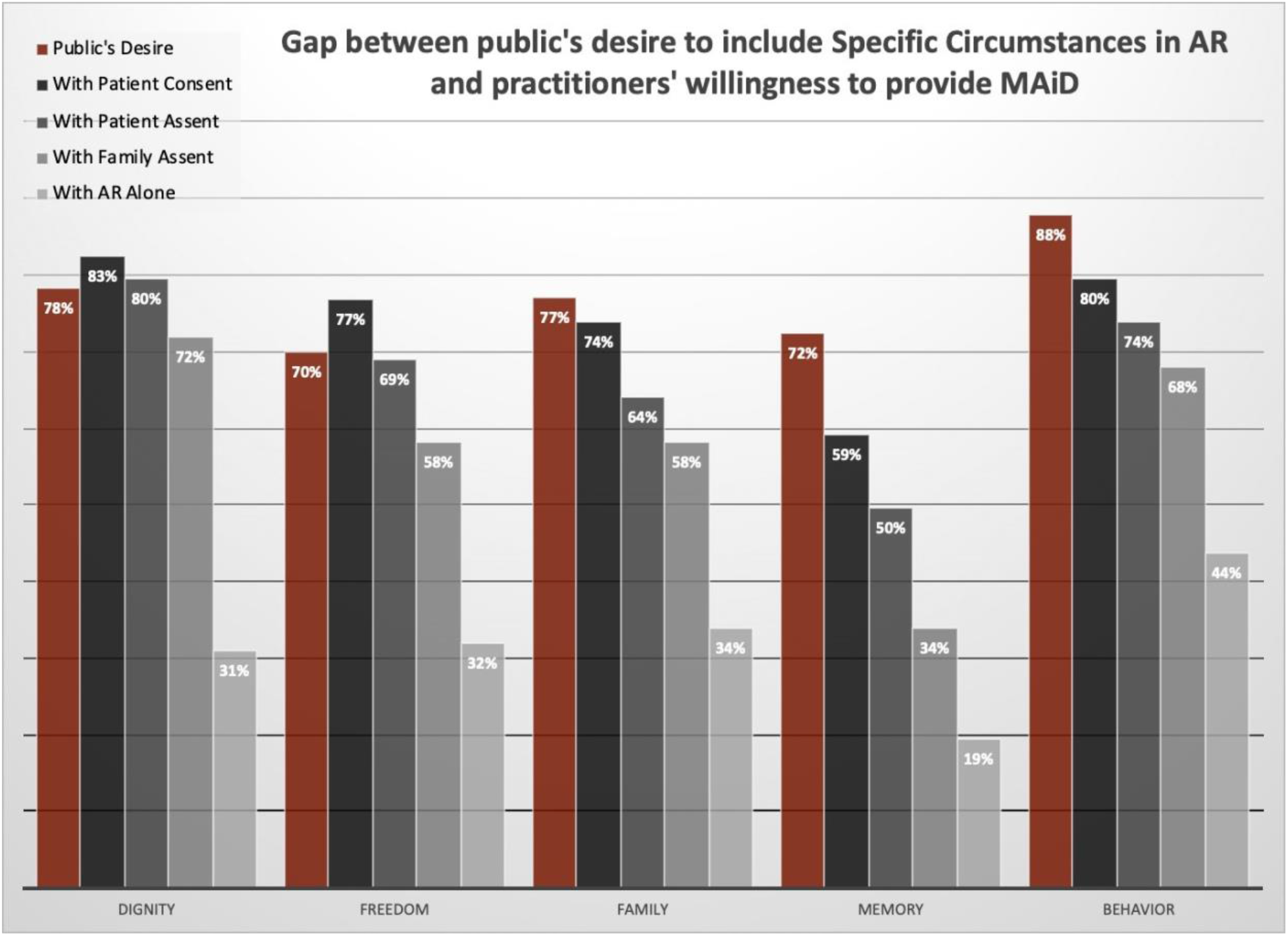
Percentage of the Canadian public who would include a given SC in their advance request, compared with the percentage of MAiD practitioners who would provide MAiD given the contextual degree of consent.

As noted in the introduction, physicians in the Netherlands rarely provide euthanasia to patients with dementia on the basis of an advance request unless there is patient assent.^5^ This represents a *gap* between what requestors anticipate will happen on the basis of their advance request and what physicians will implement. In order to explore whether a similar gap exists between the public and MAiD practitioners in Canada, we compared the likelihood that the public would include each of the 5 SCs in their advance requests with the percentage of MAiD practitioners willing to provide MAiD in that particular SC (Figure 1). We further asked MAiD practitioners whether they would be willing to provide MAiD given 4 different levels of validation by patient or family. Unsurprisingly, when patients were able to provide full consent, practitioners were as willing (or more) to provide MAiD in each SC as the public was to include it in their advance request. As levels of validation decreased through patient assent, family assent, and reliance upon the advance request alone, there was a steady increase in the magnitude of the gap between requestor desires and practitioner willingness to provide MAiD (Figure 1), consistent with the experience of dementia patients requesting euthanasia in the Netherlands.

The public were asked to comment upon why they would or would not include the various SCs in their advance requests. Common reasons for including SCs included loss of quality of life, being a burden upon family or society, and loss of self-sufficiency, while others suggested that life could still be acceptable or even enjoyable when SCs occurred (Table 3).

**Table 3:** Public’s reasons for or against including specific circumstances in an advance request

| Reasons for or against including specific circumstances in the advance request | Quote |
| --- | --- |
| Loss of quality of life | "That's basically what life is so if I can't do the normal things ..., I would want MAiD." [ID 1013] |
| Burden on family or society | "At this point I feel like I would just be a burden to employees as well as my family, and it would be easier for me to just be put out of my misery and it would be easier for those around me as well." [ID 1055] |
| Loss of self-sufficiency | "These factors are at the stage where an individual is not capable of taking care of themselves." [ID 1121] |
| Life is still acceptable | "You can still live and do productive or meaningful (in your opinion) things." [ID 1006] |

In response to the three open-ended questions, MAiD practitioners provided several hundred reasons, cautions, and operational suggestions regarding advance requests after a dementia diagnosis, taking an average of 9 minutes to answer all three free-text questions. Their reasons for why they would or would not provide MAiD were complementary to, but different than those provided by the public. Some, but not all, MAiD practitioners cited respect for precedent autonomy and acceptance of psychological suffering as a reason to provide MAiD, while specifically identifying lack of assent by patients or family, lack of overt demonstration of suffering, evidence that the patient was content, and the possibility that the advance request represents anticipatory suffering as reasons to not provide MAiD (Table 4). MAiD practitioners consistently emphasized that they would approach such cases with due respect for the individuals involved, and that providing MAiD to dementia patients would likely be emotionally challenging. Nonetheless, both the quantitative and qualitative data provide evidence that both the public and MAiD practitioners strongly support advance requests for MAiD in dementia. With respect to operational issues associated with advance requests for MAiD, practitioners emphasized that SCs should be stable over time, that regular renewal of the advance request would help practitioners at the time of MAiD, and that advance requests should detail what should transpire if things do not go as planned – i.e. if the patient resists, or family assent is not available (Table 5).

**Table 4:** Practitioners’ reasons for or against providing MAiD in response to an advance request

| Reasons for or against providing MAiD | Quote |
| --- | --- |
| Respect for precedent autonomy | "Advance request signed etc is that. We need to respect the wishes of advance consent." |
| Acceptance of psychological suffering | "I personally see the suffering and quality of life that affects people with dementia - and these people still have the right to have MAiD."<br>"It is up to the patient making the advance request to decide what would be enduring psychological suffering for them. Not up to us to challenge those values." |
| Need for patient or family assent | "I would need the person to communicate in some way that they wanted me to help them die."<br>"I think the family must be in agreement to honour their relative's advanced directives." |
| Need for evidence of suffering | "I feel capable of providing MAiD based on advance request ONLY if the patient consistently demonstrates suffering (physical or psychological) in the time period when MAiD provision is being sought. I cannot act solely on the healthy patient's anticipated suffering of their future self. If I cannot see evidence of suffering in a patient I'm asked to provide for, I cannot provide MAiD."<br>"The relief of suffering is paramount in my opinion."<br>"I am willing to respect advanced requests when suffering is apparent. I recognize suffering is personal but I have my own limits as well." |
| No MAiD if patient is content | "People's perception of psychological suffering often changes as they become demented, and sometimes they seem to be quite content with their life. Even if they have met previously determined criteria for when they want MAiD, I would not be comfortable providing MAiD for them if they appeared to be content."<br>"Truthfully if someone seemed very happy and content in their dementia with evidence they were still enjoying life (whatever that may mean), even if they met the 'bar' they had set I am not sure I would proceed." |
| Advance requests represent anticipatory suffering, not necessarily the suffering that would be experienced after onset | "In my experience as a MAiD assessor and provider, I have consistently found that what people think will be intolerable to them changes as they experience it." [ID 1047]<br>"Advanced consent remains the projection of a future that we do not know because we haven't lived it." [ID 2017] |

**Table 5:** Practitioner’s operational suggestions for implementing advance requests

| Operational suggestions for successful advance requests | Quote |
| --- | --- |
| Specific circumstances must be stable over time | "Cognition can vary widely for a patient throughout the day and between days. The other circumstances are more stable and easier to validate on my own assessment, and in collaboration with family and caregivers."<br>"I think the patient must state for how long (eg. Consistently for at least 2 months) and state something like "if there is no hope for recovery or improvement." |
| Regular renewal of advance request helps practitioners | "I would feel more comfortable with ongoing assessments with patients with dementia to confirm longitudinal and continuing wish to proceed with MAiD."<br>"I have one main concern which you did not address: how long ago was the advance directive made, and was there recent validation once the diagnosis of dementia was made and while the patient was still competent? I would have trouble offering MAiD to a patient with a remote advanced directive signed when they were much younger, just in case he or she forgot about it and would have changed their mind as they grew older."<br>"For situation where patient no longer able to participate my participation would depend on the exact details of the advance directive including how frequently & recently it was updated, whether video was included, who helped patient write it, etc. I would also want collateral information - ideally from most responsible health care provider who knew the patient prior to dementia and had discussed advance directive, etc." |
| Advance requests should detail what to do if things do not go as planned | "Requesting MAiD lucidly probably represents the biggest psychological suffering that could be. We must accept here that this suffering would still be present if not worsened in a state of confirmed dementia. Example of an AMD: "I am requesting MAiD in advance because I know well the situation that is that of patients with dementia that I am condemned to live through; and that I refuse to live through this situation, whatever may be my appearance of suffering or non-suffering when I will not be able to communicate it anymore." |

Using the method of interpretive description,^7^ these insights were abstracted and summarized into 7 policies (Table 6) which serve as recommendations for governing advance requests for MAiD after a dementia diagnosis. The underlying data was collected under the premise that a patient could individually complete an advance request for MAiD, i.e., without necessarily involving a physician. The recently passed Bill C-7 enables a form of advance request, variously termed “waiver of consent” and “advance consent”. The key difference is that Bill C-7 requires the patient and MAiD practitioner to jointly create and sign a formal document authorizing the physician to provide MAiD at some point after the patient’s loss of ability to consent. Although this form of advance consent was not under consideration when these data were collected, these recommendations are entirely relevant to the implementation of the contracts for MAiD with advance consent.

**Table 6:** Policies for governing advance requests for MAiD after a dementia diagnosis

|  |
| --- |
| **1 Formal documentation of advance request in anticipation of developing dementia**<br>Appropriate authorities in each Province should establish a standard Advance Request Form for competent residents of that Province to document their decision for the provision of MAiD once well-enumerated specific circumstances arise. This Advance Request Form should contain at least the following elements:<br>i) A list of specific circumstances for the provision of MAiD as approved by the appropriate professional society of that Province. The requestor should be required to select one or more of the specific circumstances, and may not create his or her own specific circumstance(s);<br>ii) A question asking why the selected specific circumstance(s) constitute intolerable suffering for the requestor;<br>iii) An explanation that a person’s perception of suffering may change as dementia progresses;<br>iv) A question asking whether the requestor would want to continue with MAiD if they appear to be happy or contented after the selected specific circumstance has occurred;<br>v) A question asking the requestor what they would want the MAiD practitioner to do if at the time of provision of MAiD<br>(a) the requestor does not assent or resists the procedure; or<br>(b) the requestor’s designated decision-maker or family does not assent to the procedure;<br>vi) Requirement for signature by both the requestor and legally appropriate third-party witnesses;<br>vii) Optional signature by an accredited MAiD practitioner or assessor who assisted the requestor in completing the form |
| **2 Completion of advance request form in presence of MAiD assessor**<br>Newly diagnosed dementia patients considering an advance request should be given the opportunity to meet with an accredited MAiD provider or assessor while completing the Advance Request Form. This MAiD practitioner should explain the possible specified circumstances to the requestor (and family if present) to ensure they have sufficient knowledge to properly consent, and solicit the requestor’s rationale for why the chosen circumstances constitute intolerable suffering. |
| **3 Definition of MAiD specific circumstances for advance requests in dementia**<br>The governing medical society in each Province should draw up a list of MAiD specific circumstances to be included on the Advance Request Form for that Province. These specific circumstances should be:<br>• Concrete in nature;<br>• Readily visible to practitioners, family members, and caregivers;<br>• Stable and irreversible once they have occurred; and<br>• Requiring minimal interpretive judgment by the attending physician. |
| **4 Requirement for practitioners to document and share notes about advance requests**<br>Practitioners witnessing an original advance request or a subsequent affirmation of an advance request should make contemporaneous notes of the intent of the requestor, and should provide a copy of those notes to any MAiD practitioner who is subsequently asked to provide MAiD for that requestor. |
| **5 Discretion to delay provision of MAiD if suffering can be relieved**<br>Practitioners should have the discretion to delay the provision of MAiD if they believe the patient's suffering, as described by the requestor in their advance request, could be relieved by a change in treatment or living situation, with best efforts being made to effect such a change within a reasonable amount of time. |
| **6 Periodic reaffirmation of advance request**<br>Requestors who have completed an Advance Request Form should be encouraged to meet with their MAiD practitioner or family physician on a regular basis to reaffirm their desire for MAiD upon the occurrence of any of the chosen special circumstances. |
| **7 Discussion of advance request with healthcare proxy and family**<br>Requestors should be encouraged to discuss the contents of their advance requests and the reasons for their choices with their designated healthcare decision-makers and with their immediate families, both when the advance requests are originally completed, and periodically thereafter when they are reaffirmed |

## Interpretation

### 1. Summary of results

A substantial majority of both the public and MAiD practitioner participants support advance requests for MAiD in dementia. Nonetheless, analysis of data from both the public and MAiD practitioners suggest that substantive implementation challenges remain, both in terms of the design of advance requests and contextual circumstances (i.e. levels of validation) that may be present at the time of administering MAiD. Given that Bill C-7 allows for advance consent, these issues ought to be carefully considered as part of the implementation of this Bill in order to ensure that the desires of individuals completing an advance consent are realized, while minimizing ambiguity for MAiD practitioners.

### 2. Comparison with other findings

Our finding that 86% of public respondents want the option of an advance request for MAiD after a dementia diagnosis is consistent with the results of two large trans-Canadian public polls.^8,9^ Bravo et al. found that over 70% of older adults, dementia caregivers, nurses, and physicians in Quebec agreed that it was acceptable to provide MAiD to an incompetent patient who was showing signs of distress and had requested MAiD in writing prior to losing capacity.^10^ Studies by Nuhn et al.^11^ and by Selby et al.^12^ found that the primary reasons given by patients seeking MAiD were loss of function and loss of dignity, similar to our findings reported in Table 3. The Council of Canadian Academies (CCA) Expert Panel examining advance requests concurred with at least two of the policy conclusions emerging from this study: the need for clear definition of intolerable suffering by the patient; and having the requestor describe their motivations for choosing MAiD.^13^

### 3. Future directions

The present study was carried out before Bill C-7 became law, and while the recommendations for advance requests are relevant to advance consent, future studies should explore the degree to which MAiD practitioners and other stakeholders find them to be appropriate to this new reality. Indeed, all of the proposed policies should be tested with legislators, medical societies, MAiD practitioners, legal societies, attorneys specializing in wills and advance healthcare directives (notaries in Quebec), public interest groups, and members of the public.

### 4. Limitations

An important limitation of this study is that both the public and MAiD practitioners were asked to respond to a hypothetical scenario with which they have no direct experience; what they would do when confronted with the reality of a diagnosis or a patient is unknown. The term ‘assent’ was not defined for practitioners; future studies should query MAiD practitioners using an explicit definition. The quantitative data of public responses derive from a convenience sample, and future studies should seek to test these questions on a representative sample of Canadians. While the responses from practitioners derive from a self-selected sample, it is worth noting that participants represent more than of a third of active MAiD practitioners in Canada.

### 5. Conclusion

We have identified the extent and nature of the gap between patient desires and MAiD practitioners’ willingness to provide MAID in the context of an advance request; a substantial impetus of this gap appears to be differing perspectives on the validity of anticipatory suffering. We have developed a set of recommendations designed to ameliorate the gap in order to enhance the likelihood that each person’s wishes are realized. Key among these factors are identifying specific circumstances that are stable and easily identified, clarity about what constitutes suffering to the requestor, regular re-endorsement of the advance request, inclusion of family/surrogate decision-maker in developing the advance request, and clarity as to what should be done if parties disagree. Given that advance consent is now allowed by law, consideration of these recommendations is more urgent than ever.

## Data Availability

The data that support the findings of this study are available from the corresponding author upon reasonable request.

## Funding statement

The authors received no financial support for the research, authorship, and/or publication of this article.

## Notes

### Competing Interest Statement

The authors have declared no competing interest.

### Funding Statement

The authors received no specific funding for this work.

### Author Declarations

This study was approved by the University of British Columbia Research Ethics Board (H20-02437).

